# Ongoing and planned Randomized Controlled Trials of AI in medicine: An analysis of Clinicaltrials.gov registration data

**DOI:** 10.1101/2024.07.09.24310133

**Authors:** Mattia Andreoletti, Berkay Senkalfa, Alessandro Blasimme

**Affiliations:** Department of Health Sciences and Technology, ETH Zurich, Zurich, Swtizerland

## Abstract

The integration of Artificial Intelligence (AI) technologies into clinical practice holds significant promise for revolutionizing healthcare. However, the realization of this potential requires rigorous evaluation and validation of AI applications to ensure their safety, efficacy, and clinical significance. Despite increasing awareness of the need for robust testing, the majority of AI-related Randomized Controlled Trials (RCTs) so far have exhibited notable limitations, impeding the generalizability and proper integration of their findings into clinical settings. To understand whether the field is progressing towards more robust testing, we conducted an analysis of the registration data of ongoing and planned RCTs of AI in medicine available in the Clinicaltrials.gov database. Our analysis highlights several key trends and challenges. Effectively addressing these challenges is essential for advancing the field of medical AI and ensuring its successful integration into clinical practice.

## Introduction

Machine learning (ML), Large Language Models (LLMs), and other cutting-edge technologies at the core of Artificial Intelligence (AI) hold promise to revolutionize clinical care. The potential of AI applications in the clinical space depends on their capacity to streamline patient care, to reduce medical errors, to improve diagnostic and prognostic accuracy, and to predict which therapeutic options are best suited to the specific characteristics of each patient.

While AI-specific challenges, such as limited interpretability and bias risks have been widely discussed, especially in relation to its clinical use^1^, a lesser degree of attention has been paid to issues related to the evidence base supporting the implementation of AI-based applications in the clinical context. Recently, the importance of rigorous testing and real-world validation, including well-designed Randomized Controlled Trials (RCTs), has been emphasized before AI can be integrated into medical practice. However, the vast majority (85.9%) of FDA-approved clinical AI applications (both medical devices and medical software) received 510(k) clearance^2,3^, meaning they were cleared for medical use based on tests demonstrating substantial equivalence to a predicate, that is, an already licensed product. Predicate equivalence mostly relies on non-clinical testing, especially in cases of devices that regulators classify as low-risk^4^.

Moreover, so far, a significant portion of the evidence surrounding medical AI tools has focused on their performance metrics, such as diagnostic accuracy, with dubious quality^5^. However, improved performance does not necessarily lead to a net clinical benefit for patients^6–8^. Lacking adequate clinical validation, there remains a significant gap in our understanding of the actual clinical significance of such new tools.

In recent years, the medical and scientific communities have increasingly emphasized the need for rigorous evaluation and validation of AI technologies before their integration into clinical practice. Numerous studies and commentaries have highlighted the potential risks of prematurely adopting AI-driven solutions in healthcare^9–13^. Concerns about reliability, safety, and ethical implications have dominated discussions, leading to calls for systematic evaluation frameworks and robust testing protocols^14^.

This paper aims to examine the ongoing and planned RCTs assessing medical use of AI, based on Clinicaltrials.gov registration data, and focusing on their reported characteristics. Our analysis will provide valuable insights into the current state of clinical research in medical AI.

## Methods

For our study, we queried the ClinicalTrials.gov database for studies listing ‘Artificial Intelligence’ as an intervention or treatment. The data was downloaded as an CSV file and imported into Excel to facilitate review and analysis. All available information was included in the downloaded dataset. We then filtered the results by study type to select only interventional studies, and by date to identify studies with completion dates extending beyond the search date (February 27, 2024).

Our inclusion criteria encompassed only randomized studies enabling the integration of AI into clinical practice, thereby influencing patient health management by clinical teams, as well as those permitting the application of AI-assisted tools in clinical settings, encompassing diagnosis, treatment, and prognostication.

Then, from an initial pool of 253 trials, we excluded 24 due to non-randomized status and 76 for not being applicable to our criteria. Furthermore, 21 studies lacked sufficient relevance to the medical field, and 5 failed to demonstrate clear utilization or impact assessment of AI. The remaining 127 trials comprised our final dataset for analysis [Figure 3].

We systematically extracted data from all included trials, capturing details such as whether they were multior singlecenter, conducted on a national or multinational scale, participant demographics including age and sex, medical specialties and oncological subspecialties, types of interventions employed, countries involved, enrollment status and numbers, study duration, sponsor types, and characteristics of primary outcome measures. To enhance reproducibility and maintain control over results, we minimized interpretation to a significant extent. Our analysis and subsequent visualization of the data were conducted using a combination of GraphPad Prism, MS Excel, and Adobe Photoshop.

## Results

### Study Centers and Sponsors

Of the included trials, 56.69% (n=72 of 127) were conducted in a single-center setting, while the share of multi-center clinical trials stood at 26.77% (n=34 of 127). The remaining 16.54% (n=21 of 127) did not specify the single or multi-center character of the study in the database. Only 7.87% of the included trials (n=10) were conducted in a multinational setting, while 75.59% were carried out in a single country (n=96). The remaining 16.54% (n=21) did not report country information. Beside the 21 trials that do not report study locations, the country with the highest number of trials (n=29) was the USA; followed by China with 11 trials, and Italy with 10 trials. Furthermore, Taiwan was involved in 8 studies, while France, Germany, and Spain were each involved in 7, followed closely by the UK with 5 studies and Hong Kong with 4 studies. For the rest, 24 separate countries across different continents were involved with study numbers ranging from 1 to 3.

Tangentially, looking at the sponsors of individual studies, the highest share was found to be sponsored by universities, public institutions that include hospitals and local governments, or foundations typically focusing on advancement of health, with 85.04% of the analyzed trials (n=108) falling into this category. Another 6.30% (n=8) were sponsored by industry entities like private companies and incorporations. 5.51% of the studies (n=7) were registered as sponsored by individuals, which were often observed to be medical doctors. Finally, 3.15% (n=4) of the included studies were sponsored by what could be defined as private clinics and health institutions that function under nonprofit organizational definitions [Figure 1].

**Figure 1:**
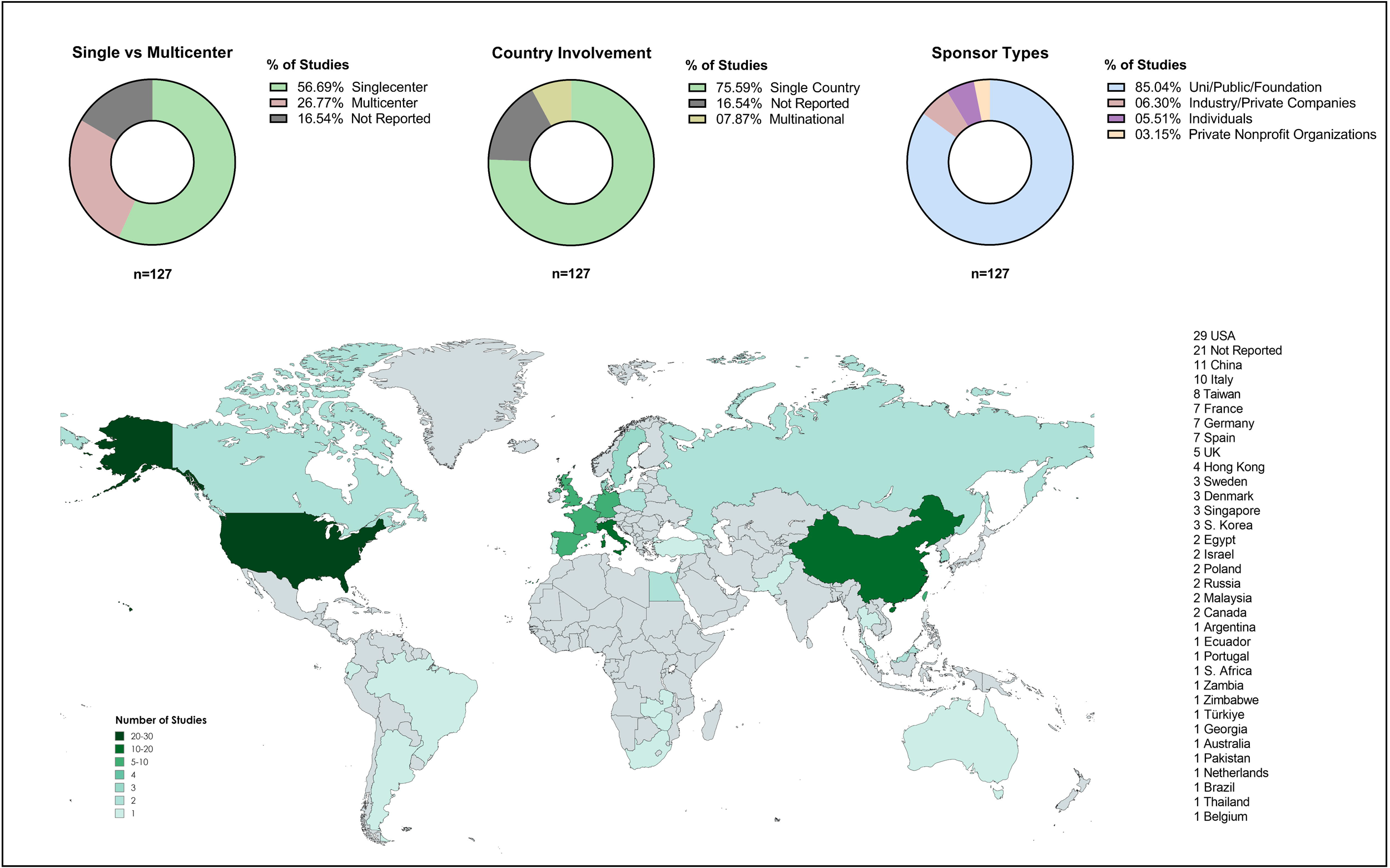
An overview of study centers and sponsor types of included studies. Included 127 studies were analyzed according to their single-versus multi-center character, single-versus multi-national design, and sponsor types. An overview of countries involved per each unique study is provided on a map of the world. For multinational studies, each country involved is registered as a unique involvement for that country.

### Demographics and Enrollment

Among the included 127 studies, 86.61% (n=110) enrolled only adults, while 9.45% (n=12) carried out the study in both children and adults, whereas only 3.94% (n=5) enrolled children only. A vast majority of the studies did not have a single-sex character, with 88.98% (n=113) enrolling both males and females. The remaining 11.02% (n=14) of the studies were sex-specific, with 9.45% (n=12) of the assessed RCTs enrolling only females, and 1.57% (n=2) enrolling males only [Figure 2a].

**Figure 2:**
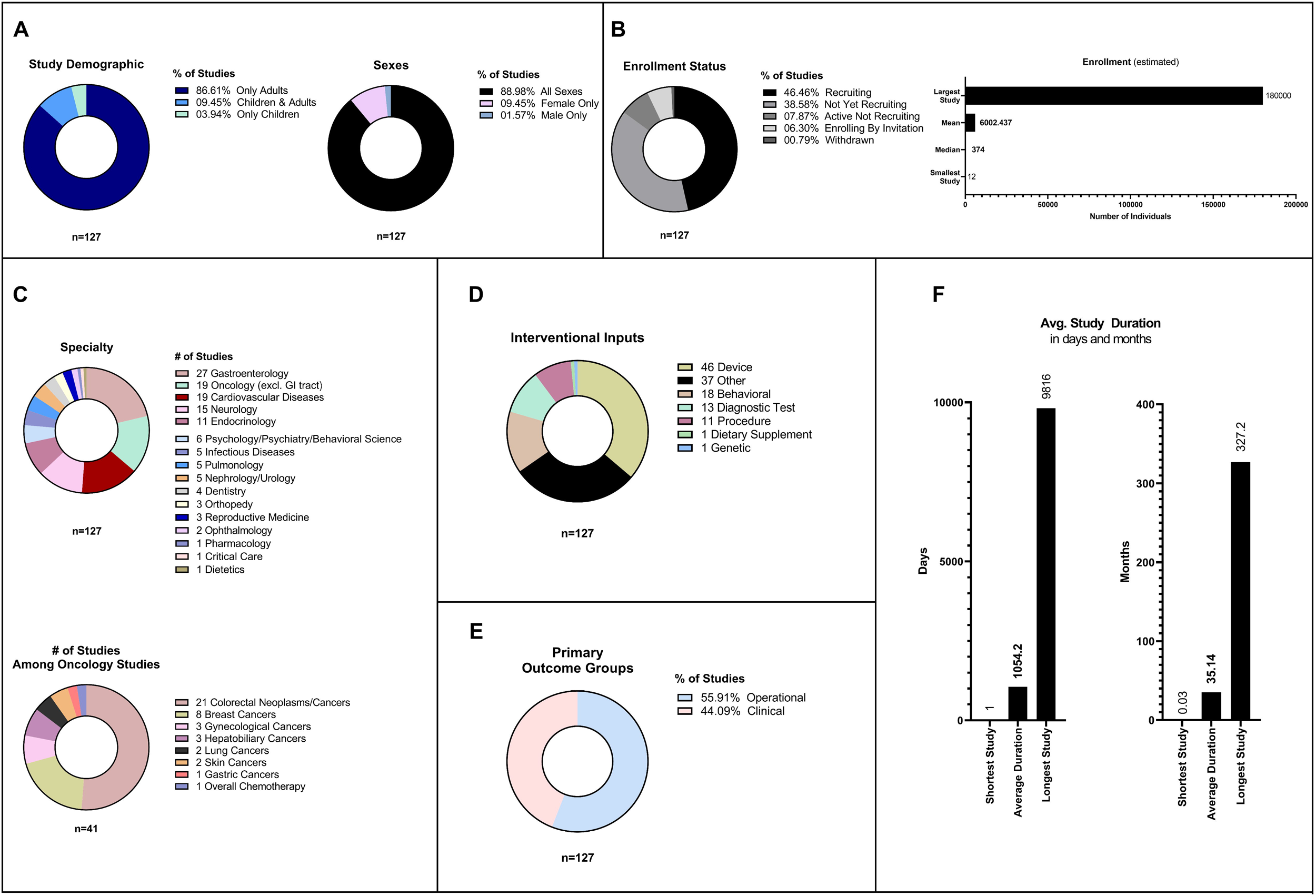
Comprehensive characteristic analyses of included studies. Comprehensive characteristic analyses of the included studies showing (A) study demographic and sexes targeted, (B) enrollment status and estimated enrollment numbers, (C) medical specialties and subspecialties of oncological studies, (D) interventional input categories, (E) primary outcome groups, and (F) average study duration. One study withdrawn before completion is not included in estimated enrollment numbers but included in the rest of the characteristic analyses. One study with the same start and completion date has been included as 1 day in the calculation of average duration data.

**Figure 3:**
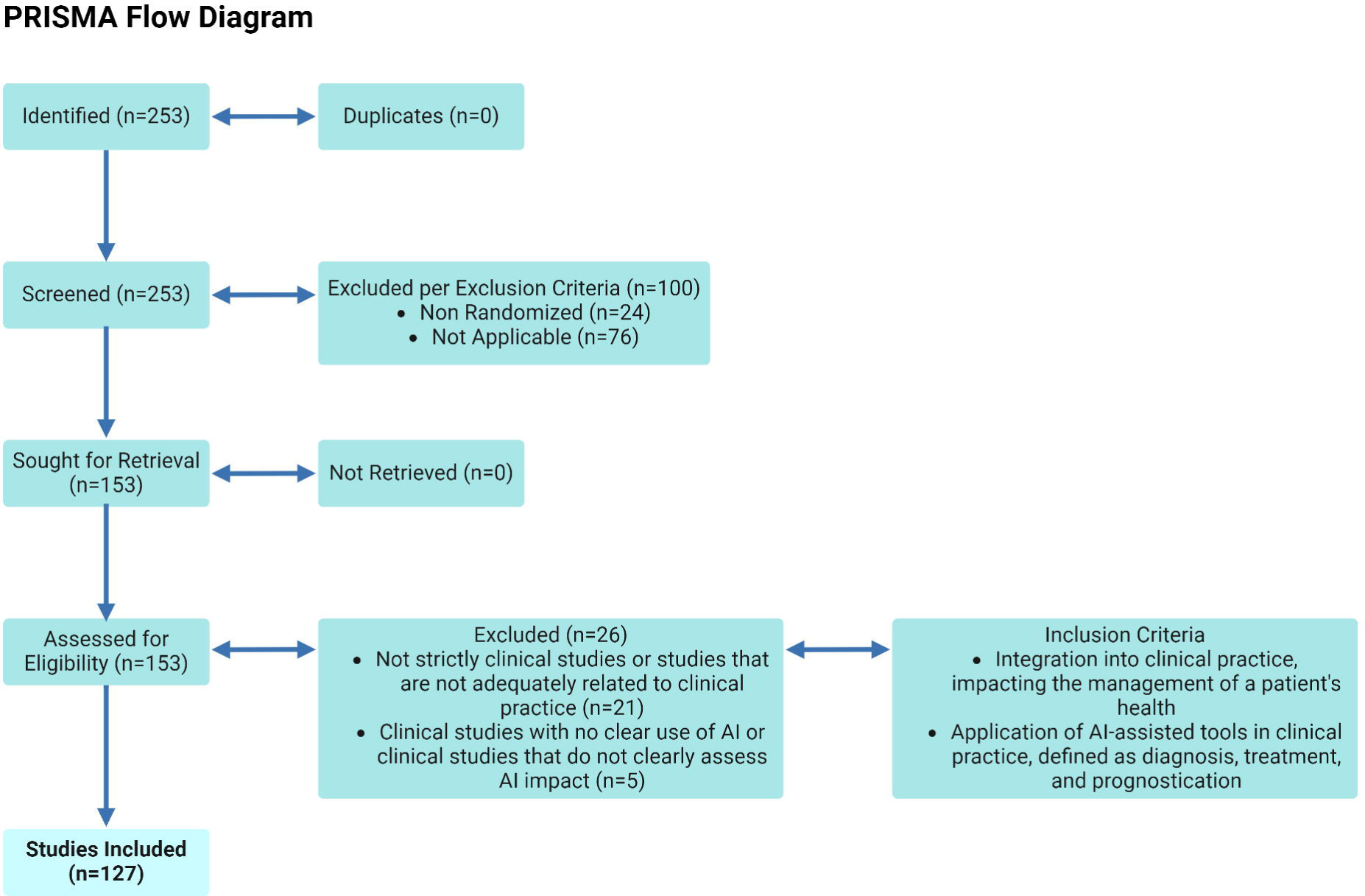
PRISMA flow diagram showing the study design and included studies. A total of 253 studies were initially identified with no duplicates. During screening, clinical trials that were non-randomized, or randomization status was given as not applicable were excluded. Among the 153 studies sought for retrieval, all were assessed for eligibility, were 26 studies that did not fully suffice the inclusion criteria were excluded, resulting in a total of 127 studies included in the subsequent analyses.

Focusing on the enrollment characteristics of the 127 evaluated studies and looking at the enrollment status, 46.46% (n=59) of the included studies were readily recruiting participants at the time of data extraction. On the other hand, 38.58% (n=49) of the RCTs were not yet recruiting study participants. Among the remaining studies, 7.87% (n=10) had ‘active, not recruiting’ status, while 6.30% (n=8) were enrolling by invitation. Finally, 1 study corresponding to 0.79% was withdrawn before completion, due to lack of funding. As for estimated enrollment numbers, excluding the withdrawn study which reported 0 participants, the trial with the highest enrollment reported 180000 participants while the study with the least enrollment reported only 12 participants. Again, excluding the withdrawn study from the 127 trials included, the mean estimated enrollment number was found to be 6002.437, while the median value stood at 374 [Figure 2b].

### Medical Specialties

The most represented medical specialty in our sample is gastroenterology (n=27), including neoplasms and cancers. This was followed by oncology excluding cancers of the gastrointestinal tract with 19 studies, and cardiovascular diseases, also with 19 studies. Neurology also had a significant presence among studies assessing AI use with 15 RCTs, followed by endocrinology with 11 studies. The remaining specialties included psychology/psychiatry/behavioral science (6 studies), infectious diseases and infectiology (5 studies), pulmonology (5 studies), nephrology/urology (5 studies), dentistry (4 studies), orthopedy (3 studies), reproductive medicine (3 studies), ophthalmology (2 studies), and pharmacology, critical care, and dietetics with 1 study for each category.

Among all 41 RCTs with an oncological focus across specialties, 21 studies were reserved for colorectal neoplasms and cancers, followed by 8 breast cancer studies, 3 studies focusing on gynecological cancers and 3 studies on hepatobiliary cancers. Among remaining studies, 2 RCTs tackled AI use in lung cancers, 2 in skin cancer, followed by 1 in gastric cancer and 1 looking at chemotherapy effects in different tumor types [Figure 2c].

### Intervention Classes

Regarding the types of interventions under testing, 46 studies focused on medical devices incorporating AI, followed by 18 studies on AI-powered behavioral interventions. Additionally, 11 studies each explored AI-assisted medical procedures and AI-based diagnostic tests. Notably, the remaining 36 studies reported various ‘other’ interventions. These diverse applications of AI in medical practice include adopting existing AI tools for improving medical practice, including for example, large language model (LLM) chatbots for diagnosis assistance, AI-based screenings, and AI apps to drive public health interventions. Due to their variety, these interventions cannot be neatly classified into predefined categories [Figure 2d]

### Primary Outcome Measures

Following established definitions, clinical outcomes are those endpoints that represent objective measurements of direct patient benefit, including metrics related to mortality, survival, and disease-associated quality of life. On the other hand, operational outcomes encompass measures such as diagnostic accuracy, procedure satisfaction, and patient anxiety. In our sample, 55.91% of the RCTs (n=71) measured operational primary outcomes, while the remaining 44.09% (n=56) focused on primary clinical outcomes [Figure 2e].

### Study Duration

Among the 127 studies assessed, the earliest study start date was recorded as March 1, 2017, while the latest study start date was set to be February 15, 2025. On the other hand, the earliest study completion date was March 1, 2024, whereas the latest study completion date was set to be June 30, 2050. Looking at study duration for all the included studies, the shortest duration recorded was 1 day (0.03 months), followed by a study of 90 days (3 months), while the longest study was set to last 9816 days (327.2 months). The average study duration for all included studies was calculated as 1054.2 days, corresponding to 35.14 months [Figure 2f].

## Discussion

Analyzing ongoing and planned RCTs of AI in medicine can offer valuable insights into whether the field is advancing towards the adoption of rigorous clinical testing standards. Our analysis provides key indications of the trajectory of clinical evaluation within this domain.

Despite repeated calls for rigorous evaluation through multicenter and multinational studies ^15^, our findings suggest that such study designs are still lacking. The majority of trials in our dataset were conducted in single-center settings, with only a small percentage being multinational. Additionally, it is noteworthy that most of these studies are concentrated in the United States and China. While not unexpected, this geographical skew may limit the generalizability of findings to other regions across the globe. These findings highlight the need for more collaborative efforts across institutions and countries to ensure the generalizability and applicability of AI-driven solutions across diverse demographics, clinical settings, and cultures.

Moreover, the studies exhibited a broad spectrum of sample sizes. This significant variance may reflect a lack of standardized approaches to sample size calculation in RCTs involving AI. The study duration also expressed high variability, nonetheless the average duration seems to be aligned with RCTs in other fields. RCTs are indeed notoriously long experiments with the median time from starting until completing follow-up reported as 2.6 years^16^.

The dominance of trials sponsored by universities and public institutions raises questions about the balance between profit-driven research and public interest. Publicly funded trials provide an opportunity for critical product optimization before entry into clinical practice by limiting commercial and market pressures in RCTs, which may otherwise bias private sponsors towards overlooking underperforming aspects or potentially harmful safety considerations of products being assessed^17^. In contrast, profit-driven RCTs may provide resources for larger-scale trials with more extensive participant recruitment and longer follow-up periods. This can enhance the statistical power and robustness of study findings, thereby improving the reliability of research outcomes. Additionally, private sponsors may have the infrastructure and expertise to conduct complex multicenter or multinational trials, which can provide more diverse and representative findings. One potential rationale for this phenomenon could be the dynamic and intricate nature of the regulatory framework surrounding AI in healthcare, which is still in the process of evolution. Uncertainty about regulatory requirements, including approval processes and compliance standards, may deter companies from investing in RCTs for AI-driven medical interventions. The risk of regulatory hurdles or delays could discourage private-sector sponsors from undertaking clinical trials. Moreover, companies may be hesitant to invest in profit-driven RCTs for AI applications if there is uncertainty about reimbursement policies, or healthcare provider adoption rates. On the one hand, the lack of clear pathways to commercialization and monetization could further discourage companies from allocating resources to conducting well-designed trials. On the other hand, competition in a fast-growing market may create incentive to favor the most rapid way to regulatory clearance – that is by showing predicate equivalence (510k clearance) instead of seeking clinical validation via RCTs.

The analysis of medical specialties targeted by AI-focused trials provides insight into areas where AI is expected to have the greatest impact. Gastroenterology, oncology, and cardiovascular diseases emerged as the primary areas of focus, indicating a forthcoming trend in these domains. An interesting dissection can be made that a significant portion of the RCTs assessing medical devices, with 21 of the 46 studies falling under the gastroenterology specialty, showing an apparent trend for AI use in general, and medical devices incorporating AI in particular for gastroenterology practice. Among all the observed types of interventions being tested, medical devices being the most prevalent intervention class highlights a narrow focus within the realm of AI applications, despite the potential for a much broader range of uses^18^.

The predominance of operational primary outcome measures, particularly those related to diagnostic accuracy and precision, underscores the emphasis on improving clinical workflows and decision-making processes. While operational outcomes are essential for assessing the practical implications of AI tools, it is crucial to also prioritize clinical endpoints that directly impact patient outcomes, such as mortality, survival rates, and quality of life, to better capture the actual effectiveness and clinical significance of AI interventions in healthcare.

Notably, our findings align closely with the results of reviews on published RCTs of medical AI ^19–23^. These reviews consistently highlight several prevalent challenges, which we also observed in our sample. Most trials were single-center studies, focused on operational endpoints rather than clinically significant outcomes, and were based on small samples. These features significantly hinder the assessment of medical AI and its integration into clinical practice. However, a limitation of these reviews is that the field of AI, including medical AI, is evolving very rapidly. As a result, reviews that rely on published RCTs, which were designed long before their results were published, cannot capture the latest trends. Our analysis of ongoing and planned trials indicates that this trend has not significantly changed. As noted, there is a pressing need for more internationally collaborative, multi-center trials with larger sample sizes, diverse outcome measures, and clinically meaningful endpoints.

However, it is important to also acknowledge that conducting RCTs for AI applications in medicine is inherently complex and faces numerous methodological and practical challenges^24,25^. These obstacles, coupled with the elevated costs associated with RCTs, the need to swiftly penetrate the market, the absence of clear regulatory incentives, and the lack of a standardized regulatory framework for medical AI applications, are likely forestalling the development of more robust testing standards. RCTs for testing medical AI are still in their early stages. The challenges highlighted in our review, as well as in other studies, should be meticulously addressed in the design of future trials.

It is important to acknowledge the limitations of our study. We queried only one database (Clinicaltrials.gov) to maintain methodological uniformity, as different databases use varying classification parameters. Additionally, the potential for bias due to the lack of pre-registration of trials must be recognized. This omission could unintentionally skew our data analysis and interpretation.

## Data Availability

All data produced in the present study are available upon request to the authors.

## Data availability

All data produced in the present study are available upon request to the authors.

## Author contributions

MA*: Designed the study, collected the data, and drafted the manuscript.

SB*: Organized and analyzed the data, designed the figures, and contributed to drafting the manuscript.

AB: Contributed to the study design and revised the manuscript.

^*^Equally contributed to the work.

## Competing Interests

The authors declare no competing interests.

## Acknowledgements

This study did not receive any funding.

## Notes

### Competing Interest Statement

The authors have declared no competing interest.

